# Proteome-wide analysis of differentially-expressed SARS-CoV-2 antibodies in early COVID-19 infection

**DOI:** 10.1101/2020.04.14.20064535

**Authors:** Xiaomei Zhang, Xian Wu, Dan Wang, Minya Lu, Xin Hou, Hongye Wang, Te Liang, Jiayu Dai, Hu Duan, Yingchun Xu, Yongzhe Li, Xiaobo Yu

## Abstract

Rapid and accurate tests that detect IgM and IgG antibodies to SARS-CoV-2 proteins are essential in slowing the spread of COVID-19 by identifying patients who are infected with COVID-19. Using a SARS-CoV-2 proteome microarray developed in our lab, we comprehensively profiled both IgM and IgG antibodies in forty patients with early-stage COVID-19, influenza, or non-influenza who had similar symptoms. The results revealed that the SARS-CoV-2 N protein is not an ideal biomarker for COVID-19 diagnosis because of its low immunogenicity, thus tests that rely on this marker alone will have a high false negative rate. Our data further suggest that the S protein subunit 1 receptor binding domain (S1-RBD) might be the optimal antigen for IgM antibody detection, while the S protein extracellular domain (S1+S2ECD) would be the optimal antigen for both IgM and IgG antibody detection. Notably, the combination of all IgM and IgG biomarkers can identify 87% and 73.3% COVID-19 patients, respectively. Finally, the COVID-19-specific antibodies are significantly correlated with the clinical indices of viral infection and acute myocardial injury (p≤0.05). Our data may help understand the function of anti-SARS-CoV-2 antibodies and improve serology tests for rapid COVID-19 screening.

---

Since December 2019, coronavirus disease 2019 (COVID-19) has become a worldwide pandemic. As of April 14, 2020, COVID-19 has spread to 185 countries with 1,929,922 confirmed cases and 120,450 deaths. COVID-19 symptoms range from a mild cough to pneumonia, and it is estimated that ~17.9% of patients have mild symptoms or no symptoms at all (*1–4*). This subgroup of patients, although contagious, would not be selected for viral RNA testing and quarantined. However, diagnostic testing is critical to effective containment of SARS-CoV-2, which is the virus responsible for COVID-19 (*5*).

SARS-CoV-2 antibodies are produced within the first week of infection (*6, 7*). Thus, tests to detect SARS-CoV-2 antibodies in serum, plasma, and whole blood have been developed to rapidly screen patients to ascertain infection status and possibly immunity to COVID-19 (*8*). Current rapid antibody tests from different manufacturers rely on antibody binding to SARS-CoV-2 nucleocapsid (N) protein, spike (S) protein, or S protein fragments (i.e., subunit 1, S1; receptor binding domain, RBD). Both N and S proteins are structural proteins. Some manufacturers even use a combination of the N and S proteins and protein fragments, thus resulting in varying or inconsistent antibody tests (*8, 9*). The SARS-CoV-2 genome also encodes a polyprotein (the open reading frame 1a and 1b, Orf1ab), two additional structural proteins (envelope, E; membrane, M) and five accessary proteins (Orf3a, Orf6, Orf7a, Orf8, Orf10)(*10*). The development of a rapid *in vitro* diagnostic serology test with high sensitivity and specificity relies on selecting the antigens with high immunogenicity. Furthermore, it important to consider the homology of the antigen with proteins from other viruses to decrease the number of false negative results (*8, 11*).

In this study, we used a SARS-CoV-2 proteome microarray developed in our laboratory to perform a proteome-wide analysis of differential antibody responses to SARS-CoV-2 proteins in the serum of 40 patients displaying similar symptoms (i.e., fever, cough or muscle ache) with COVID-19, influenza, or non-influenza (Figure 1a, Table 1, Supplementary Figure 1)(*12*). The early-stage COVID-19 patients {Onset of symptoms, 4.0 (1.0-20.0) days} were diagnosed according to the Diagnosis and Management Plan of Pneumonia with New Coronavirus Infection (trial version 7). The non-influenza controls are the patients with similar symptoms as COVID-19, but excluded the infection of respiratory virus (Table 1).

**Figure 1.**
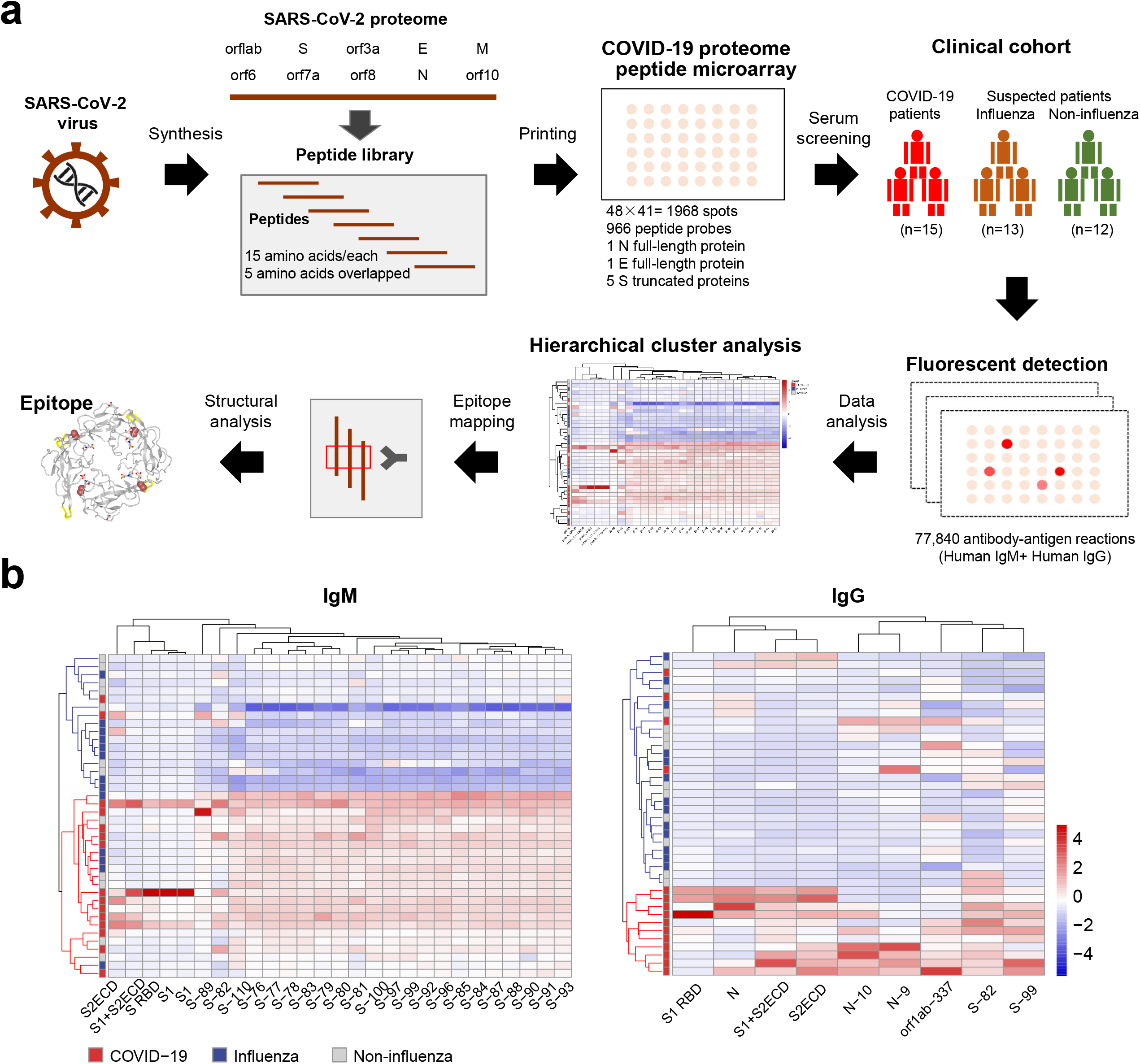
Proteome-wide analysis of differential antibody response to SARS-CoV-2 proteins using a SARS-CoV-2 proteome microarray. (a) The workflow of proteome-wide analysis of differential antibody response to SARS-CoV-2 proteins using a proteome microarray. (b) Hierarchical clustering analysis of differential antibody responses to SARS-CoV-2 proteome between patients with COVID-19, influenza, and non-influenza displaying similar symptoms. The false-colored rainbow color from blue to red correspond to low to high signal intensities on the array, respectively.

**Table 1.** Clinical characteristics of early COVID-19 and suspected patients displaying similar symptoms

|  | **Non-influenza**<br>**patients**<br>**( n=12 )** | **Influenza patients**<br>**(n=13)** | **Covid-19 patients**<br>**(n=15)** |
| --- | --- | --- | --- |
| **Demographics and clinical characteristics** |  |  |  |
| male | 3 (25%) | 8 (61.5%) | 7 (46.7%) |
| female | 9 (75%) | 5 (38.5%) | 8 (53.3%) |
| Age, year | 36.5 (32.0-57.0) | 51.0 (20.0-88.0) | 38.0 (7.0-68.0) |
| 2019-nCoV(+) | 0 | 0 | 15 (100%) |
| FluA-RNA(+) | 0 | 6 (46.2%) | 0 |
| FluB RNA(+) | 0 | 3 (23%) | 0 |
| RSV RNA(+) | 0 | 4 (30.8%) | 0 |
| Exposure history | 2 (16.7%) | 4 (30.8%) | 14 (93.3%) |
| Fever | 12 (100%) | 13 (100%) | 11 (73.3%) |
| Headache | 3 (25%) | 4 (30.8%) | 1 (6.7%) |
| Cough | 4 (33.3%) | 6 (46.2%) | 7 (46.7%) |
| Sputum | 3 (25%) | 3 (23%) | 2 (13.3%) |
| Myalgia | 1 (8.3%) | 7 (53.8%) | 2 (13.3%) |
| Fatigue | 1 (8.3%) | 6 (46.2%) | 0 |
| Diarrhea | 0 | 1 (7.7%) | 0 |
| Dyspnea | 0 | 1 (7.7%) | 2 (13.3%) |
| Nausea or vomiting | 0 | 0 | 0 |
| Onset of symptoms, days | 1.0 (0-5.0) | 2.0 (1.0-22.0) | 4.0 (1.0-20.0) |
| **Imaging features** |  |  |  |
| Ground-glass opacity | 8 (66.7%) | 7(53.8%) | 12 (80%) |
| Bilateral pulmonary infiltration | 4 (33.3%) | 6(46.2%) | 10 (66.7%) |

Data are median (IQR) or n (%).

The SARS-CoV-2 proteome microarray contained full-length N protein, full-length E protein, five S truncated proteins and 966 tiled peptides representing the SARS-CoV-2 proteome (Supplementary Table 1). Each peptide was 15 amino acids long with a 5 amino acid overlap. The intra- and inter-array correlation of serum IgM and IgG antibody detection were 0.9916 to 0.9992, respectively, demonstrating the high-reproducibility of the SARS-CoV-2 proteome microarray (Supplementary Figure 2). Using the microarray, we analyzed the serum of 15 COVID-19 confirmed patients, 13 influenza patients and 12 non-influenza patients, which generated a total of 77,840 antibody-antigen reactions that were screened.

First, we analyzed the elevated antibodies in early-stage COVID-19 patients compared to the influenza and non-influenza patients (Supplementary Figure 1). A comparison of the COVID-19 and non-influenza patient groups identified 76 IgM antigens and 17 IgG antigens that were differentially targeted (Supplementary Figure 3). The IgM antigens included the S protein (26 peptides, S1, S1RBD, S1+S2ECD, S2ECD), N protein (1 peptide, full-length protein), M protein (2 peptides), E protein (3 peptides, full-length protein), Orf1ab (10 peptides), Orf3a (18 peptides), Orf6 (5 peptides), Orf8 (1 peptide), and Orf10 (3 peptides). The IgG antigens included the S protein (5 peptides, S1, S1RBD, S1+S2ECD, S2ECD), N protein (2 peptides, full-length protein), E protein (1 peptides), and Orf1ab (4 peptides). A comparison of the COVID-19 and influenza groups identified 86 IgM antigens and 64 IgG antigens (Supplementary Figure 4). The IgM antigens included the S protein (67 peptides, S1, S1RBD, S1+S2ECD, S2ECD), N protein (2 peptides), M protein (2 peptides), and Orf1ab (10 peptides). The IgG antigens included the S protein (10 peptides, S1RBD, S1+S2ECD, S2ECD), N protein (2 peptides, full-length protein), and Orf1ab (48 peptides) (Supplementary Table 2).

**Table 2.**
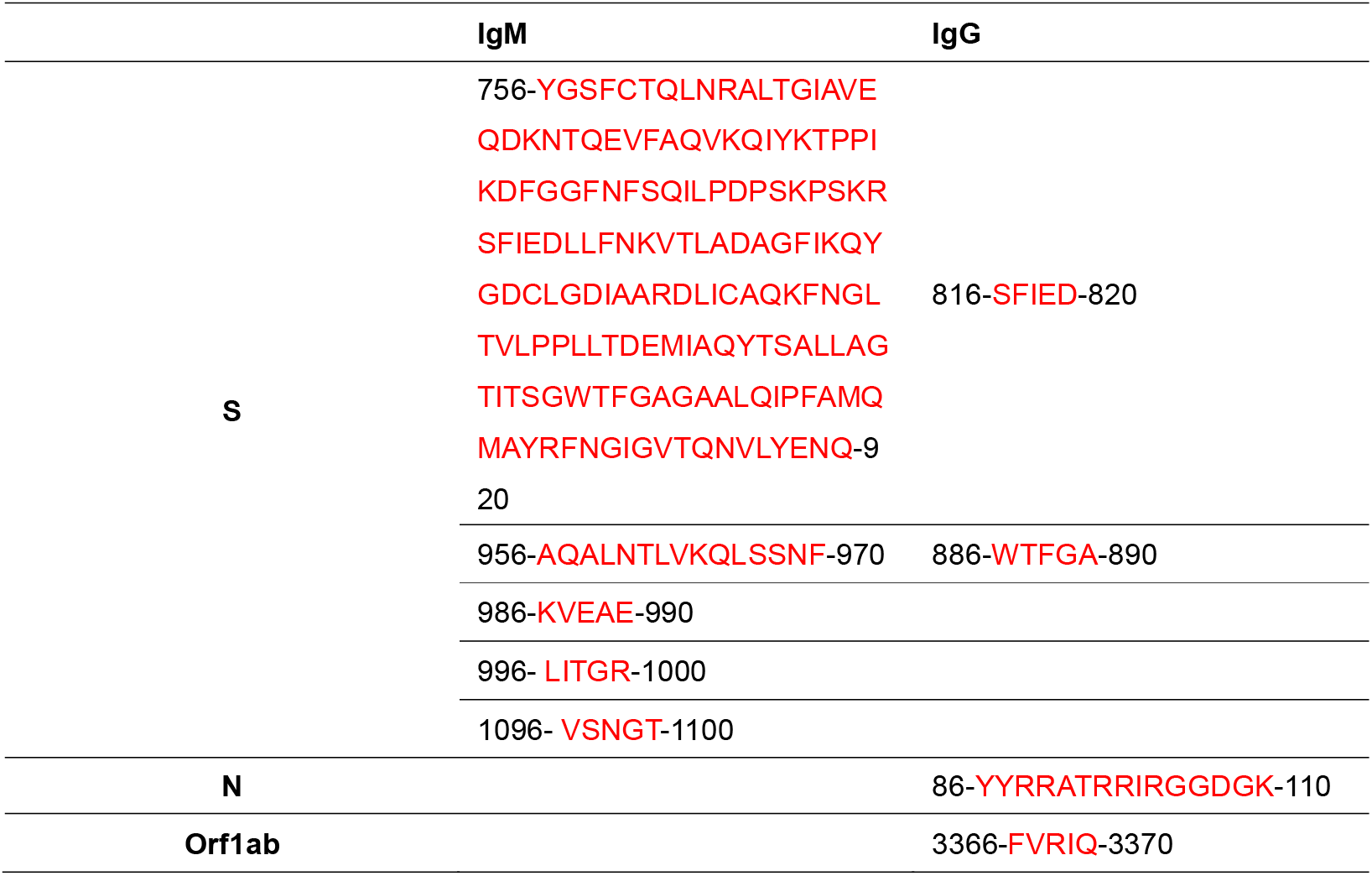
Binding epitopes of anti-SARS-CoV-2 antibodies

A Venn diagram analysis indicates that 27 IgM antigens and 9 IgG antigens were unique to COVID-19 compared to both influenza and non-influenza groups (Supplementary Figure 5). All of the IgM antigens were from the S protein (22 peptides, S1, S1RBD, S1+S2ECD, S2ECD). The IgG antigens were from the S protein (2 peptides, S1RBD, S1+S2ECD, S2ECD), N protein (2 peptides, full-length protein) and Orf1ab (1 peptide) (Supplementary Table 2). These results suggest that, during the early stage of COVID-19 infection, IgM antibodies targeting the S protein are produced. The IgG anti-SARS-CoV-2 antibodies, which are produced later during COVID-19 infection, bind to the S, N and Orf1ab proteins. These results make sense because the S protein is displayed across the virion surface, which is easily accessible to the humoral immune system. The N and Orf1ab proteins, on the other hand, is usually compartmentalized within the viral particle. As the virus replicates within the host cells, the N and Orf1ab proteins could be released from the cells and later recognized by the humoral immune system(*13*).

Hierarchical cluster analyses show that 86.7 % (13/15) and 73.3% (11/15) of COVID-19 patients can be distinguished from influenza and non-influenza patients by measuring these differentially-expressed IgM and IgG antibodies, respectively (Figure 1b). These numbers are superior to the current serological antibody tests using Immunocolloidal Gold lateral flow, ELISA and Chemiluminescence technologies, which have positive rates (%) ranging from 7% to 53% from the same serum samples (Supplementary Figure 6). These results indicate the potential of our SARS-CoV-2 proteome microarray in identifying appropriate antibody biomarkers for early COVID-19 diagnosis.

In order to know which SARS-CoV-2 proteins are suitable for developing a diagnostic test, we compared the IgM and IgG antibodies of COVID-19, influenza, and non-influenza patients in regards to how they targeted two structural proteins, N and S (N, S1, S1RBD, S1+S2ECD and S2ECD). It is surprising that the N protein IgM antibodies cannot discriminate between COVID-19 and the other influenza and non-influenza patients. Instead, IgM antibodies targeting S1-RBD and S1+S2ECD could best discriminate COVID-19 patients from the other groups with a p-value less than 0.01 (COVID-19 vs. influenza) and 0.001 (COVID-19 vs. non-influenza) (Figure 2a). These results were validated using a peptide microarray, in which four antibody binding epitopes on the S protein subunit 2 (S2) were identified (Table 1, Supplementary Table 2). Furthermore, the structural analysis of the trimerized S protein indicate that four epitopes are located within the extracellular domain of subunit 2 (S2ECD, Ser686-Pro1213). These epitopes included 756-920 amino acid residues (YGSFCTQLNRALTGIAVEQDKNTQEVFAQVKQIYKTPPIKDFGGFNFSQILPDPSKPSKRSFIEDLLFNKVTLADAGFIKQYGDCLGDIAARDLICAQKFNGLTVLPPLLTDEMIAQYTSALLAGTITSGWTFGAGAALQIPFAMQMAYRFNGIGVTQ NVLYENQ), residues 956-970 (AQALNTLVKQLSSNF), residues 1096-1100 (VSNGT), and residues 996-1000 (LITGR) (Figure 3a).

**Figure 2.**
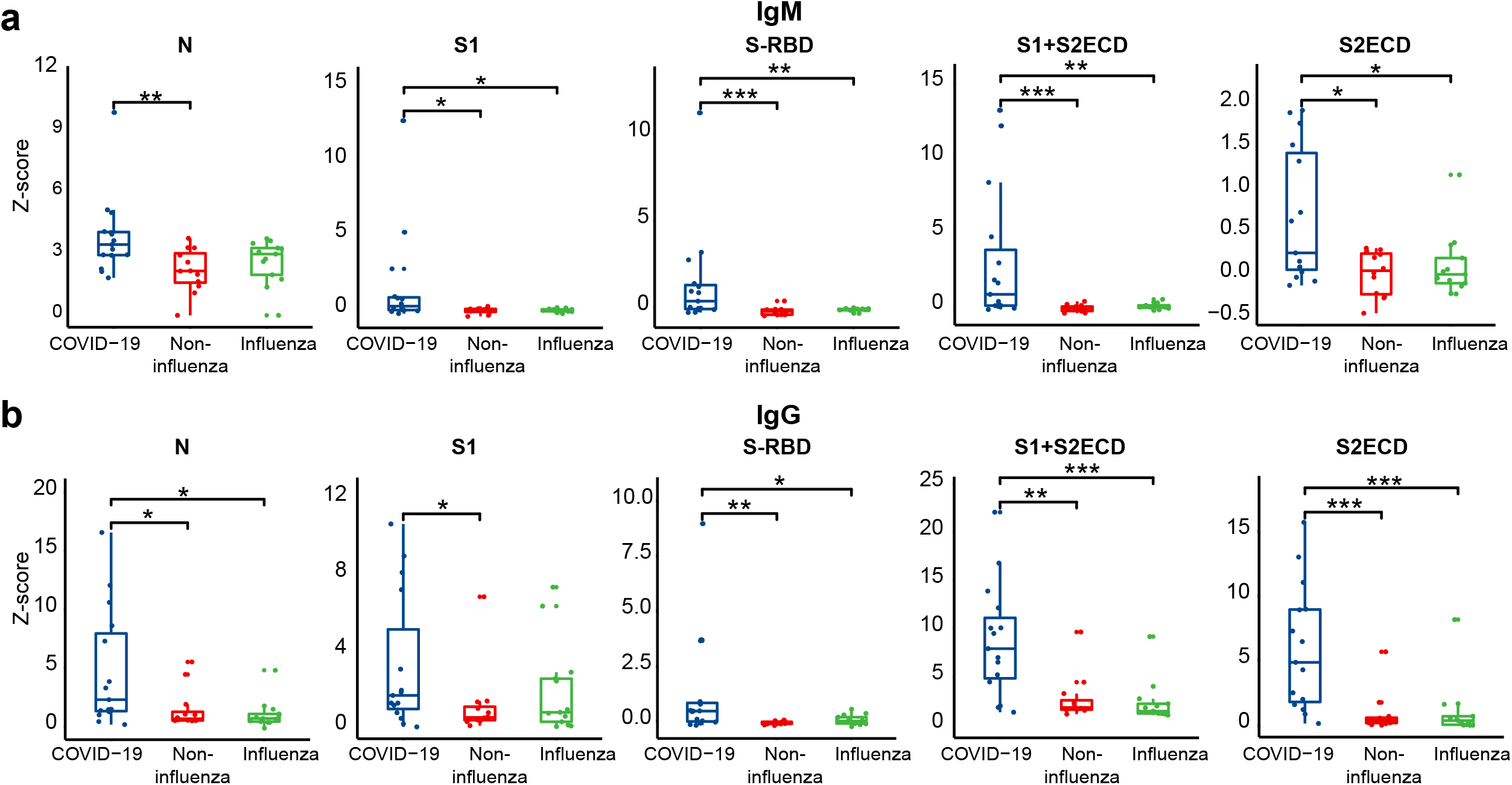
Box plot analysis of antibody responses to SARS-CoV-2 structural N and S proteins between confirmed and suspected COVID-19 patients. The statistical significance was calculated using Mann Whitney *U*-test with a p-value less than 0.05. The one, two and three asterisks represent the p-value of 0.05, 0.01 and 0.001, respectively.

**Figure 3.**
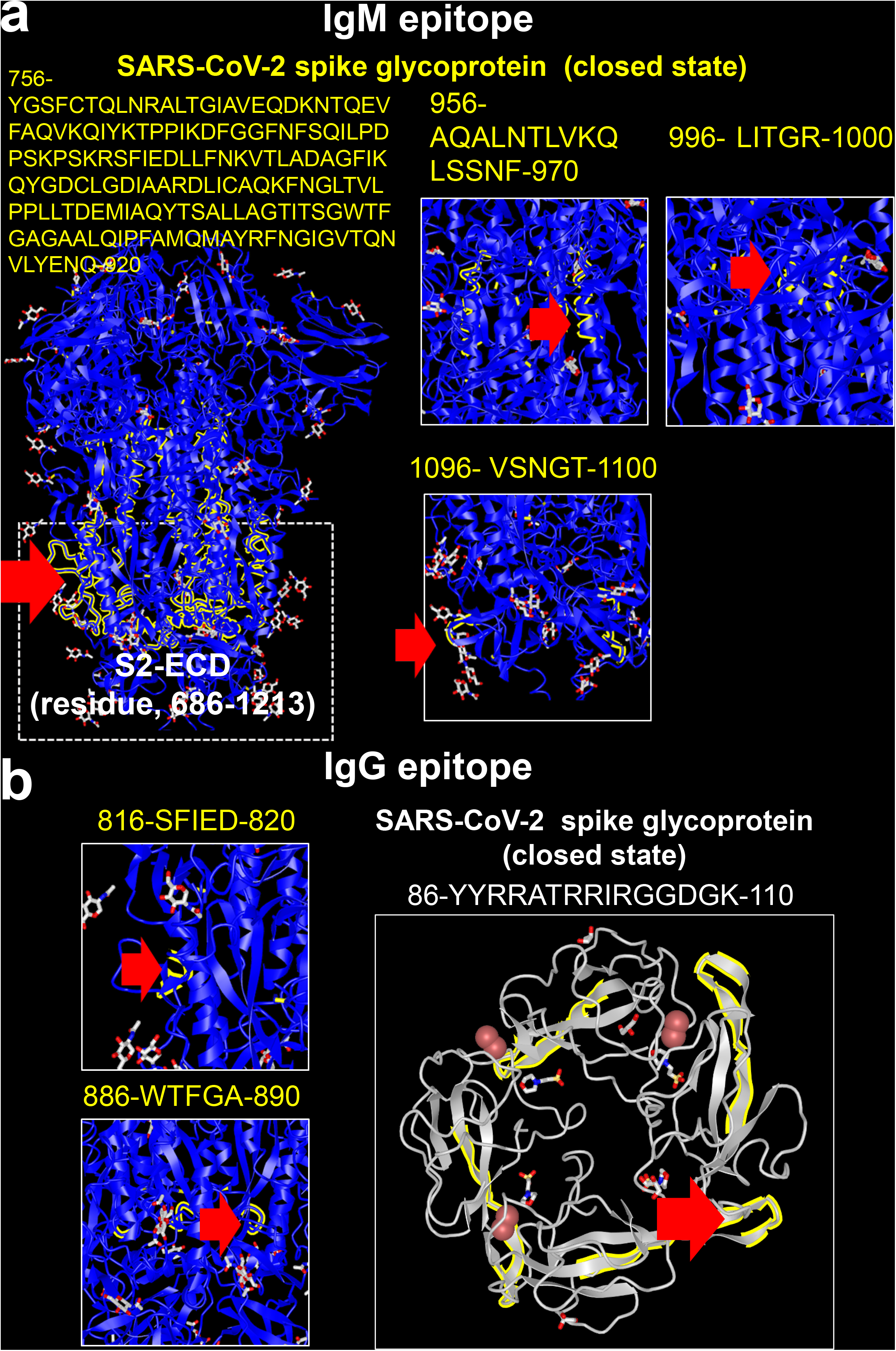
Structural analysis of immunogenic epitopes of SARS-CoV-2 proteins. Structural analyses of the (a) nucleocapsid protein’s RNA binding domain (PDB ID: 6VYO) and (b) spike protein (PDB ID: 6VXX). The epitope is labeled in yellow and indicated with a red arrow.

The concentration of IgG antibodies to N, S, S1+S2ECD, S1-RBD, and S2ECD proteins was significantly higher in COVID-19 patients than the other groups, with S2ECD having the best discriminative performance. The results were also validated using a peptide microarray, in which four peptides from N, S, and Orf1ab proteins were identified as the antibody binding epitopes (Table 2, Supplementary Table 2). Sequential alignment and structural analysis indicate that two epitopes on the S protein (residues 816-820, SFIED; residues 886-890, WTFGA) are located on the surface and inside of the S protein, respectively (Figure 3b). Notably, the one epitope (residues 86-100, YYRRATRRIRGGDGK) on the N protein is located within the RNA binding domain loop that is easily accessible to antibodies. These results suggest that using the appropriate control groups (e.g., non-COVID-19 patients with similar symptoms) is critical in selecting antibody biomarkers for an antibody-based diagnostic test (Supplementary Figures 3 and 4).

The N protein is not an ideal antigen for the diagnostic test at early SARS-CoV-2 infection because the IgM antibodies that target this protein could not discriminate between the COVID-19 patients from the influenza patients (Figure 2a). The sensitivity of IgG antibodies for N protein is also low and only 30% (5/15) early COVID-19 patients can be discriminated (Figure 2b). The finding can be confirmed by a meta-analysis of 7848 individuals, in which the sensitivity of S antigen-based tests is superior to that of N antigen-based tests (*14, 15*). The S1-RBD and S1+S2ECD proteins might be the optimal antigens for the IgM antibody test, whereas the S2ECD protein might be the optimal antigen for the IgG antibody test (Figure 2). Furthermore, the combination of different biomarkers could increase the test’s sensitivity compared to using a single biomarker alone by distinguishing 86.7 % (13/15) and 73.3% (11/15) COVID-19 patients from the influenza and non-influenza patients (Figure 1b). Identification of multiple biomarkers or peptide epitopes with high immunogenicity is possible with our SARS-CoV-2 proteome array.

Finally, we performed a comprehensive circos correlation analysis of COVID-19 specific SARS-CoV-2 antibodies and 20 clinical indices. Statistically significant correlations (p≤0.05) are indicated in pink (Figure 4, Supplementary Table 3). The IgM antibodies to the S protein significantly correlated with eosinophil count (EOS#), hemoglobin (HGB), platelet count (PLT), albumin (Alb), creatine kinase MB mass (CKMB-mass) and N-terminal pro-brain natriuretic peptide (NT-proBNP) levels. On the other hand, the IgG antibodies to the S protein significantly correlated with lymphocyte count and percentage (LY# and LY%), CKMB-mass and NT-proBNP levels. Notably, the levels of two biomarkers of cardiac injury, NT-proBNP and CKMB-mass, were significantly correlated with both IgM and IgG COVID-19 specific antibodies (Figure 4), suggesting that anti-SARS-CoV-2 antibodies contribute to acute myocardial injury(*16*).

**Figure 4.**
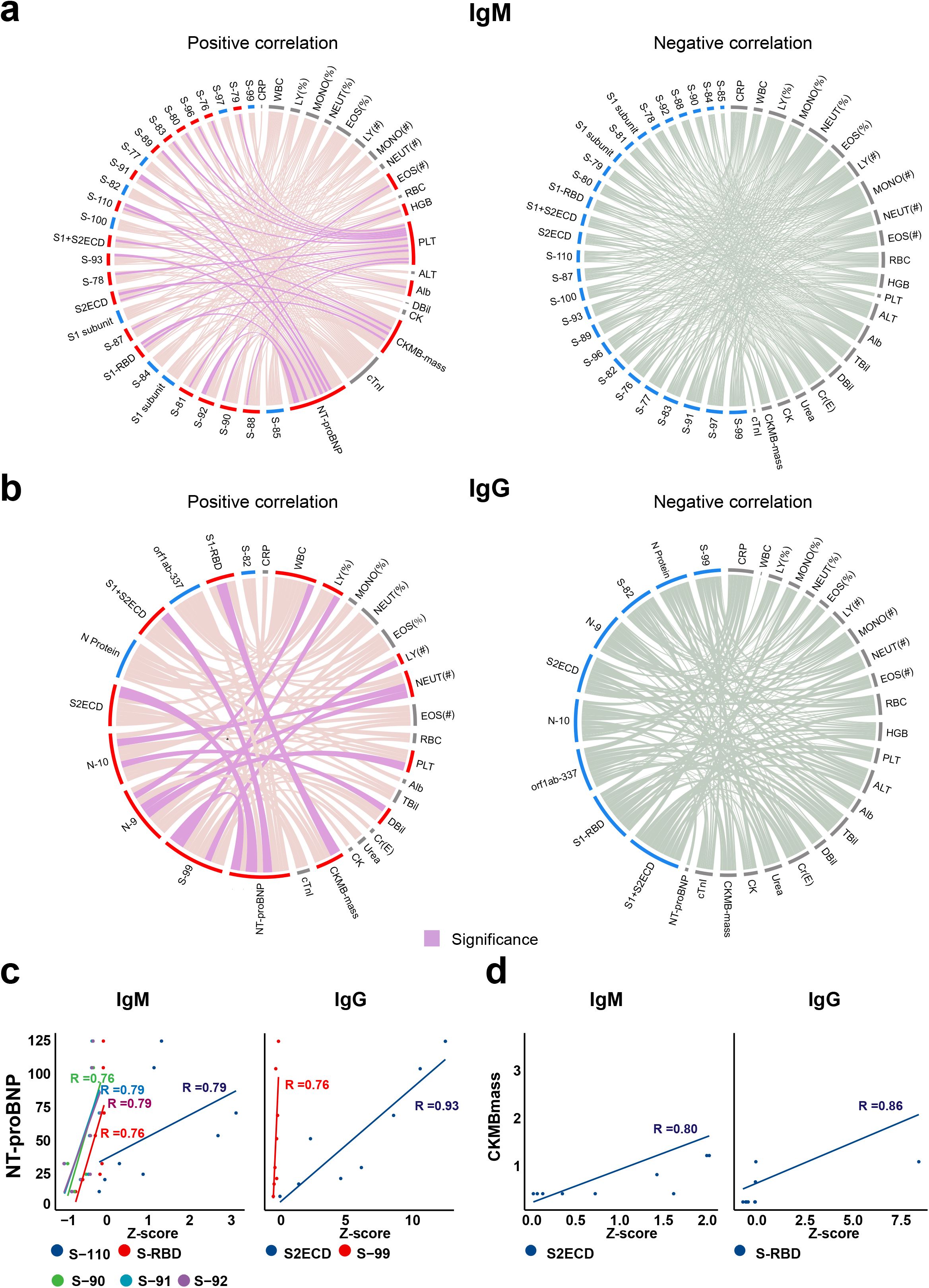
Correlation network of COVID-19 specific antibodies and clinical indices. (A) IgM and (B) IgG antibody correlations between COVID-19 specific antibodies and clinical index using circos. Correlation with statistical significance (p≤0.05) are indicated in pink. Positive and negative correlations with non-significance (p≥0.05) are indicated in red and gray, respectively. (c) and (d) are the COVID-19 antibodies correlated with NT-proBNP and CKMBmass, respectively. The COVID-19 antibodies were selected with a Spearman coefficient correlation higher than 0.6 and a p-value less than 0.05.

A limitation of this study is that antibodies binding to conformational epitopes or post translational modifications could not be detected using our microarray comprised of chemically-synthesized, linear peptides (*17*). In addition, a small number of patients were employed. These results should be validated using a large patient cohort with other respiratory infections(*16*).

Our work provides valuable information to help fight COVID-19. Understanding the humoral immune response to COVID-19 will help us understand the role of antibodies in COVID-19, identify potential targets for immunotherapy, and enable the production of accurate rapid serology tests for COVID-19 detection. Detection of IgM antibodies will help identify those with acute COVID-19 infection who need to be quarantined. Detection of IgG antibodies may help identify those who are recovering from or have gained immunity to COVID-19.

## Data Availability

The data is available upon the request of correspondering author.

## Contributions

X. H., Y. L. X. W. provided the clinical samples. X. Z., H. W., D. W., J. D. prepared the microarrays. X. H., X. W. and X. Z. executed microarray experiments. D. W., X. Z., T. L. and X.Y. executed the statistical and structural analysis. X.Y., and Y. L. conceived the idea, designed experiments, analyzed the data and wrote the manuscript.

## Acknowledgement

This work was supported by the State Key Laboratory of Proteomics (SKLP-C202001, SKLP-O201703 and SKLP-K201505), the Beijing Municipal Education Commission, National Natural Science Foundation of China (81671618, 81871302, 81673040, 31870823), the National Program on Key Basic Research Project (2018YFA0507503, 2017YFC0906703 and 2018ZX09733003) and the CAMS Initiative for Innovative Medicine (2017-I2M-3-001 and 2017-I2M-B&R-01). We also thank Dr. Brianne Petritis for her critical review and editing of this manuscript.

## Competing interests

None declared.

## Supplementary information

The supplementary information includes the materials and methods, 3 supplementary tables and 6 supplementary figures.

## Methods

### Collection of clinical samples

Patients with COVID-19, influenza or non-influenza with similar symptoms were enrolled in the Outpatient department of Peking Union Medical College Hospital. All serum samples were collected under the approval of the intuitional review board (IRB) from Peking Union Medical College Hospital (Ethical number: ZS-2303) and Beijing Proteome Research Center. Written informed consent was waived due to the rapid emergence of this infectious disease. All experiments were performed according to the standards of the Declaration of Helsinki. The COVID-19 patients with mild symptoms were diagnosed according to the Diagnosis and Management Plan of Pneumonia with New Coronavirus Infection (trial version 7).

### Screening of serological antibodies in COVID-19 and suspected patients using SARS-CoV-2 proteome microarray

The SARS-CoV-2 proteome microarrays containing 966 peptides and full-length N, full-length E and truncated S proteins were prepared as described previously(*12*). Prior to antibody detection, the microarrays were assembled in an incubation tray and blocked with 5% (w/v) milk in 1xPBS with 0.05% (v/v) Tween-20 (PBST) for 1 min at room temperature. After washing with PBST three times, the array was incubated with 1:100 diluted serum for 30 min at room temperature and washed again. The array was then incubated for 30 min with a mixture containing Cy3 Affinipure donkey anti-human IgG(H+L) and Alexa Fluor 647 Affinipure goat anti-human IgM FC5µ antibody (Jackson ImmunoResearch, USA) (2μg/mL). Finally, the resulting array was washed with PBST and water, dissembled from the tray and dried with centrifugation for 2 min at 2,000 rpm. The slide was scanned with a GenePix 4300A microarray scanner (Molecular Devices, Sunnyvale, CA, USA) at 532 and 635 nm to measure Cy3 and Alexa Fluor 647 fluorescence, respectively. Median spot intensity minus background was extracted using GenePix Pro7 software (Molecular Devices, Sunnyvale, CA, USA).

### Detection of serological antibody in early COVID-19 patients using Immunocolloidal Gold Strip, ELISA and Chemiluminescence

Serological antibodies in patients with COVID-19, influenza and non-influenza suspected patients were analyzed using Immunocolloidal Gold Strip, ELISA and Chemiluminescence according to the manufacturers’ instructions.

### Statistical analysis

All microarray signals were normalized with a Z-score prior to statistical analyses(*18*). Differentially-expressed SARS-CoV-2 antibodies were identified using Mann Whitney *U*-test with a p-value of 0.05. The hierarchical cluster analysis of serological antibody response to peptides was performed using the R pheatmap. The circos plot was made using circos (http://circos.ca/).

## Notes

### Competing Interest Statement

The authors have declared no competing interest.

### Clinical Trial

ZS-2303

